# Colistin resistance among the Gram-negative nosocomial pathogens in India: A systematic review and meta-analysis

**DOI:** 10.1101/2024.05.01.24306721

**Authors:** Sambit K. Dwibedy, Indira Padhy, Aditya K. Panda, Saswat S. Mohapatra

## Abstract

**Background:** The rapid rise of nosocomial infections and the growing ineffectiveness of frontline antibiotics against Gram-negative bacteria (GNB) have put the healthcare sector under unprecedented stress. In this scenario, colistin, an antibiotic of the polymyxin class, has become the last resort treatment option. The unrestricted use of colistin in the preceding decades has led to the emergence of colistin-resistant (Col^R^) bacterial strains. Unfortunately, comprehensive data on the prevalence of Col^R^ nosocomial pathogens in India are lacking. This study was conducted to address this information gap and to determine the prevalence of Col^R^ among the nosocomial GNB species in India.

**Methods:** A systematic review and meta-analysis were conducted to determine the prevalence of Col^R^ among the nosocomial GNB species in India and their geographical distribution. A systematic search of the online databases was performed and eligible studies meeting the inclusion criteria were used for qualitative synthesis. The combined event rate and 95% confidence interval were estimated using forest plot with a random-effect model. Cochrane Q statistics and I^2^ statistics were used to detect possible heterogeneity.

**Results:** From a total of 1865 retrieved records from 4 databases, 36 studies were included in the study. Among the most common nosocomial pathogens*, K. pneumoniae* showed a rate of Col^R^ at 17.4%, followed by *P. aeruginosa* (14.1%), *E. coli* (12.3%), and *A. baumannii* (12.2%). Interestingly, our analysis revealed that *E. cloacae* has the highest rate of Col^R^ at 28.5%.

**Conclusions:** The level of resistance displayed by *K. pneumoniae*, *P. aeruginosa*, and to a lesser extent, *E. coli* and *A. baumannii* in the Indian subcontinent poses a challenge for public health management. Though prevalence may differ among regions and over time, continued surveillance, and efforts to curb the spread of resistance are crucial to ensure the continued effectiveness of this critical antibiotic.

## 1. Introduction

The rise and spread of antibiotic-resistant bacterial pathogens, specifically of the Gram-negative type, is one of the foremost challenges in the healthcare sector. Colistin, the cationic lipopeptide antibiotic of the polymyxin class functions by targeting the outer membrane of Gram-negative bacteria (GNB) and is used as a last option antibiotic against them [1]. Colistin is also used in animal feed, and poultry farms for prophylactic reasons. However, the overuse of colistin in the past few decades both in the clinic and animal feeds has led to the rise of colistin resistance (Col^R^) among the GNB species [2]. Considering the consequences of the widespread Col^R^ in the clinical setting and the environment, the Government of India has banned the use of colistin in animal feed since 2019.

Increasing incidences of Col^R^ among the GNB species such as *Escherichia coli*, *Klebsiella pneumonia*, *Pseudomonas aeruginosa*, and *Acinetobacter baumannii* responsible for hospital-acquired (nosocomial) infections are reported from across the globe [1]. This has posed a formidable challenge in the healthcare sector due to the limited therapeutic alternatives available. Various infections caused by these pathogens such as gastroenteritis, neonatal meningitis, UTI, hemolytic-uremic syndrome, peritonitis, mastitis, pneumonia, and septicemia by *E. coli* [3]; septicemia, urinary tract infection (UTI), bloodstream infection, and pneumonia by *K. pneumoniae* [4]; cystic fibrosis, ventilator-associated pneumonia, central line-associated bloodstream infection, and catheter-related infection by *P. aeruginosa* [5]; meningitis, bacteremia, and pneumonia by *A. baumannii* [6] among the immunocompromised and hospitalized individuals are particularly troublesome from a treatment point of view. These GNB species are the primary causes of the most difficult-to-treat infections reported in India [7].

Intrinsic resistance to polymyxins is exhibited by a variety of GNB species such as *Morganella morganii*, *Proteus mirabilis*, *Aeromonas hydrophilia, Burkholderia cepacia*, *Serratia marcescens,* etc. [1]. Moreover, resistance can also arise by spontaneous mutations in the genes encoding bacterial two-component signalling systems (TCSs) involved in membrane remodelling [1]. However, a major factor driving the global rise of Col^R^ is the plasmid-mediated *mcr* (mobile colistin resistance) gene, first reported in China in 2015 [8]. After its first report, *mcr* genes belonging to 10 different classes have been detected across the globe [9].

The use of colistin though has been prohibited in the food industry, as a leading manufacturer and user of this antibiotic, increasing incidences of Col^R^ bacteria are reported from across India. Using a meta-analysis, the prevalence of resistance to polymyxins has been estimated to be 15.0% in India, which is higher than the global average [10]. Moreover, distinct patterns of distribution of such resistant strains among various provinces and Union Territories of India have also been observed [10]. However, till date, no comprehensive analysis has been done to determine the prevalence of Col^R^ among the common nosocomial GNB species in India, though several studies on individual bacterial species in clinical settings have been reported [11–13]. These reports from India are primarily part of antimicrobial surveillance programs, in which strains are randomly collected and analysed. Inadequate data on the prevalence and spread of Col^R^ pathogenic GNB strain limit the understanding of the basis of species-specific resistance and their geographic distribution pattern. The current systematic review and meta-analysis were undertaken to estimate the rate of Col^R^ among the most common GNB pathogens involved in nosocomial infections and explore their spread in the Indian subcontinent.

## 2. Materials and methods

A systematic review and meta-analysis were performed to determine the rate of Col^R^ among the most common nosocomial GNB species in India and their nationwide spread. The guidelines of Systematic Reviews and Meta-Analyses (PRISMA) were followed for the analysis [14,15].

### 2.1 Data collection

Keywords such as “colistin resistance”, “colistin-resistant *E. coli*”, “colistin-resistant *K. pneumoniae*”, “colistin-resistant *P. aeruginosa*”, or “colistin-resistant *A. baumannii*” were used to extensively to search databases such as PubMed, Google Scholar, Scopus, and Science Direct. Original articles available in English, reporting on Col^R^ GNB isolates in India, published till April 2024 (last search date: April 10, 2024) were included in this study.

### 2.2 Inclusion and exclusion criteria

Reports having data on Col^R^ isolate, isolation from India, and identification up to the species level were included for further analysis. Broth micro-dilution (BMD), the gold standard for Col^R^ detection, was selected as the resistance detection method. Studies with cases of missing data, as well as review papers, papers that did not provide original data, and studies that employed methods other than BMD for Col^R^ determination, were excluded from the analysis.

### 2.3 Data extraction

Two authors (SKD and IP) individually assessed each potential study for eligibility after retrieving the full article. Disagreements were cleared by consulting a third author (AKP). Following a comprehensive analysis, the author’s name, publication year, country, state, total isolates studied, number of Col^R^ isolates, isolate source, resistance detection technique, and bacteria identification method were extracted and summarized from each article by two authors.

### 2.4 Meta-analysis

Comprehensive Meta-Analysis (CMA) software (version 4.0; Biostat Inc. USA) was used for the statistical analysis. The combined event rate and 95% confidence interval were estimated using a forest plot. Cochrane Q statistics [16] and I^2^ statistics [17] were used to detect possible heterogeneity. Based on the result of the heterogeneity analysis, a fixed-effect model (I^2^<50%; homogenous) was used for pooled analysis of resistance rate, or a random-effect model (I^2^>50%; heterogeneous) was used. Additionally, publication bias was assessed using funnel plots [18] and Egger’s regression analysis [19]. Sensitivity analysis was performed to explore the robustness of the meta-analysis.

## 3. Results

### 3.1 Literature search

Out of a total of 1865 retrieved records from 4 databases (PubMed = 877, Google Scholar = 543, Science Direct = 277, Scopus = 168), 36 studies through multiple evaluations were included in the qualitative analysis following the eligibility criteria (Supplementary Figure 1). Among the total 16,347 GNB isolates, 1759 were found to be Col^R^. From the total retrieved GNB isolates, 7350 were *K. pneumoniae* from 24 studies, 3476 *E. coli* isolates from 17 studies, 3266 *A. baumannii* from 13 studies, 1553 *P. aeruginosa* from 9 studies, and 702 *E. cloacae* from 4 studies were subjected to meta-analysis.

### 3.2 Publication bias and heterogeneity

The funnel plots deciphering the studies of Col^R^ among different GNB species did not suggest publication bias based on visual inspection (Figure 1). Egger’s regression analysis also supported the absence of publication bias in the analysis of the rate of Col^R^ among *E. coli* (intercept: 0.84, 95% CI = -2.37 to 4.04, p = 0.59), *K. pneumoniae* (intercept: -0.58, 95% CI = -4.37 to 3.21, p = 0.75), *P. aeruginosa* (intercept: -0.20, 95% CI = -3.8 to 3.41, p = 0.90), and *A. baumannii* (intercept: 0.53, 95% CI = -2.19 to 3.25, p = 0.68) isolates in India (Table 1). However, in the case of *E. cloacae* an asymmetrical funnel plot with a higher concentration of studies on the right side was observed (Figure 1E), though Egger’s regression analysis suggested the absence of publication bias (intercept: 4.5, 95% CI = -3.76 to 12.75, p = 0.14) (Table 1). Cochrane Q and I^2^ measurements demonstrated significant heterogeneity among the included studies (Table 1). Hence, a random-effects model was used for all meta-analyses.

**Figure 1.**
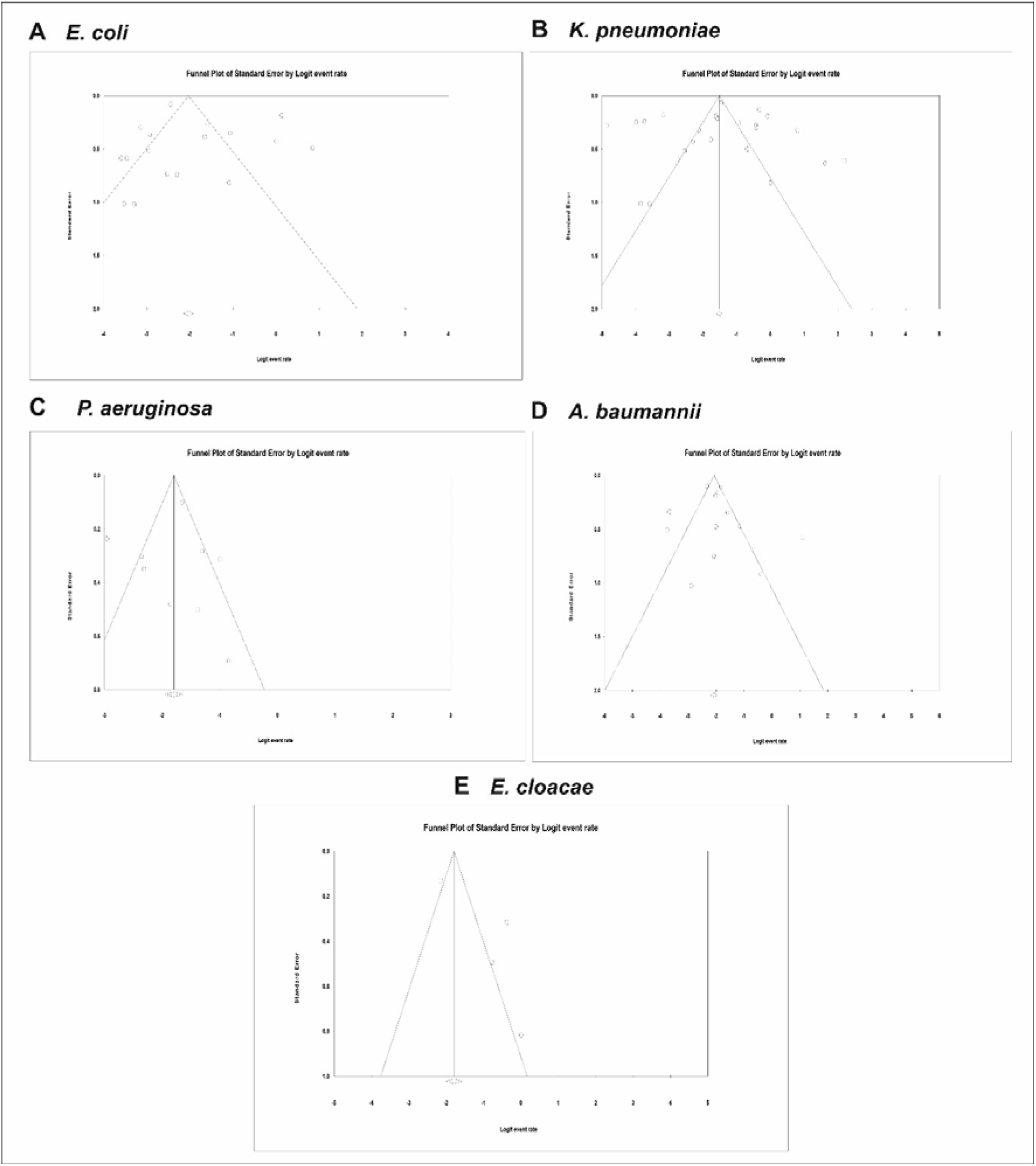
Funnel plots representing publication bias for the bacterial strains. (A) *E. coli*, (B) *K. pneumoniae*, (C) *P. aeruginosa*, (D) *A. baumannii*, and (E) *E. cloacae*.

**Table 1.** Statistics to test the publication bias, heterogeneity, used model of meta-analysis, and pooled Col^R^ prevalence.

| Study Name<br>(Incidence of Col<sup>R</sup><br>GNB) | Egger's Regression Analysis |  |  | Heterogeneity Analysis |  |  | Model<br>Used | Pooled<br>Prevalence |
| --- | --- | --- | --- | --- | --- | --- | --- | --- |
|  | Intercept | 95% Confidence<br>Interval | P Value<br>(2 tailed) | Q value | P-Heterogeneity | I<sup>2</sup> (%) |  |  |
| *K. pneumoniae* | -0.58 | -4.37 to 3.21 | 0.75 | 740.38 | 0.00 | 96.76 | Random | 17.4 |
| *E. coli* | 0.84 | -2.37 to 4.04 | 0.59 | 275.2 | 0.00 | 94.19 | Random | 12.3 |
| *P. aeruginosa* | -0.20 | -3.8 to 3.41 | 0.90 | 43.95 | 0.00 | 81.8 | Random | 14.1 |
| *A. baumannii* | 0.53 | -2.19 to 3.25 | 0.68 | 85.55 | 0.00 | 85.97 | Random | 12.2 |
| *E. cloacae* | 4.5 | -3.76 to 12.75 | 0.14 | 36.85 | 0.00 | 91.86 | Random | 28.5 |

### 3.3 Pooled Col^R^ rate among different GNB isolates

The study primarily aimed to determine the prevalence of Col^R^ among the most common nosocomial GNB pathogens. Our analysis revealed that the pooled Col^R^ rate for the pathogens were as follows; *E. coli* (341 out of 3476 isolates), *K. pneumoniae* (793 out of 7350 isolates), *P. aeruginosa* (200 out of 1553 isolates), *A. baumannii* (337 out of 3440 isolates), and for *E. cloacae* (92 out of 702) respectively. The pooled Col^R^ prevalence rate for the individual bacterial species is presented as forest plots in Figures 2 and 3.

**Figure 2.**
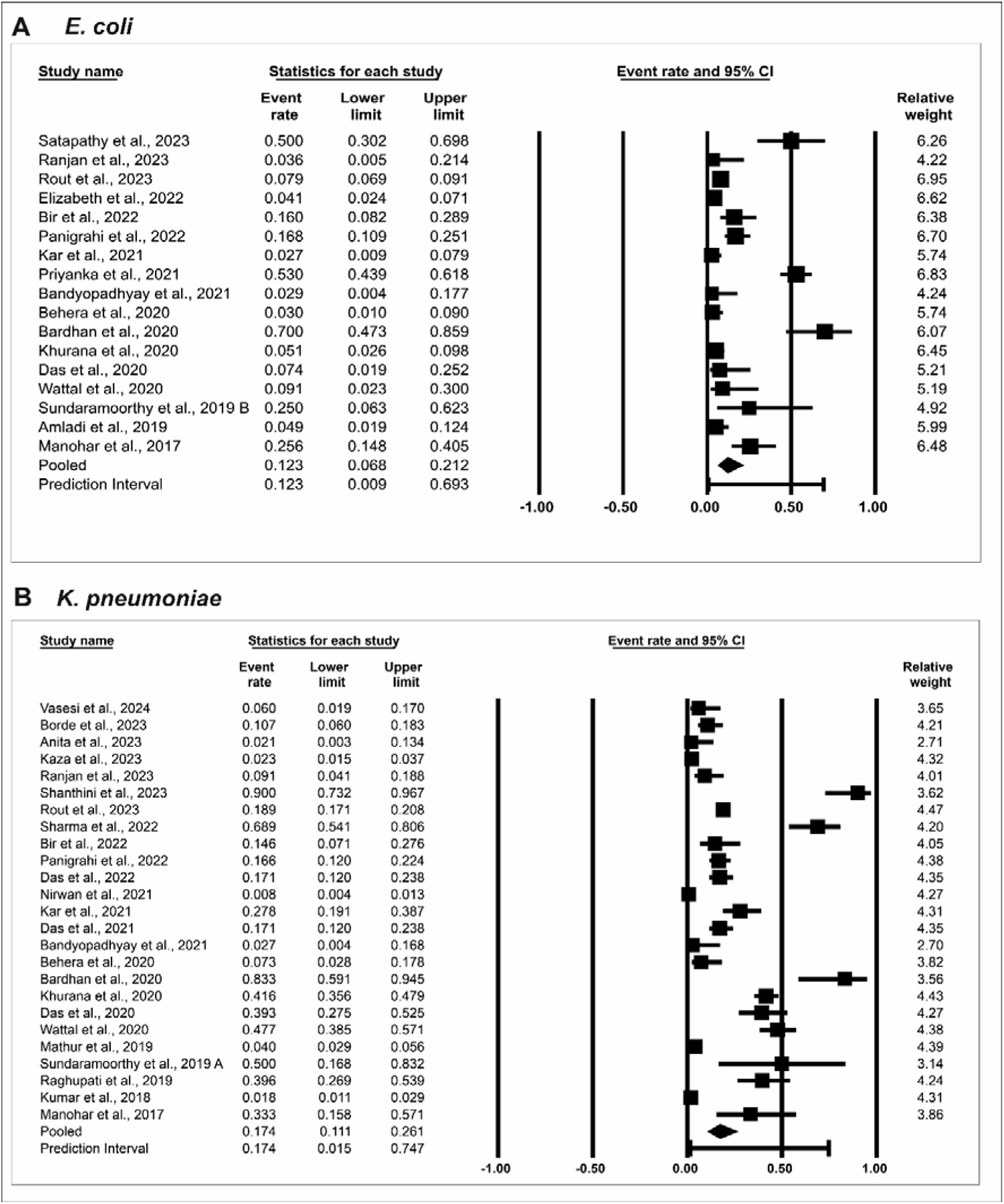
Forest plots demonstrating the pooled prevalence of Col^R^ among (A) *E. coli*, and (B) *K. pneumoniae*. The Comprehensive meta-analysis software V4 (Biostat Inc. USA) was used for the calculation of the event rate, and 95% confidence interval, and used in the forest plot.

**Figure 3.**
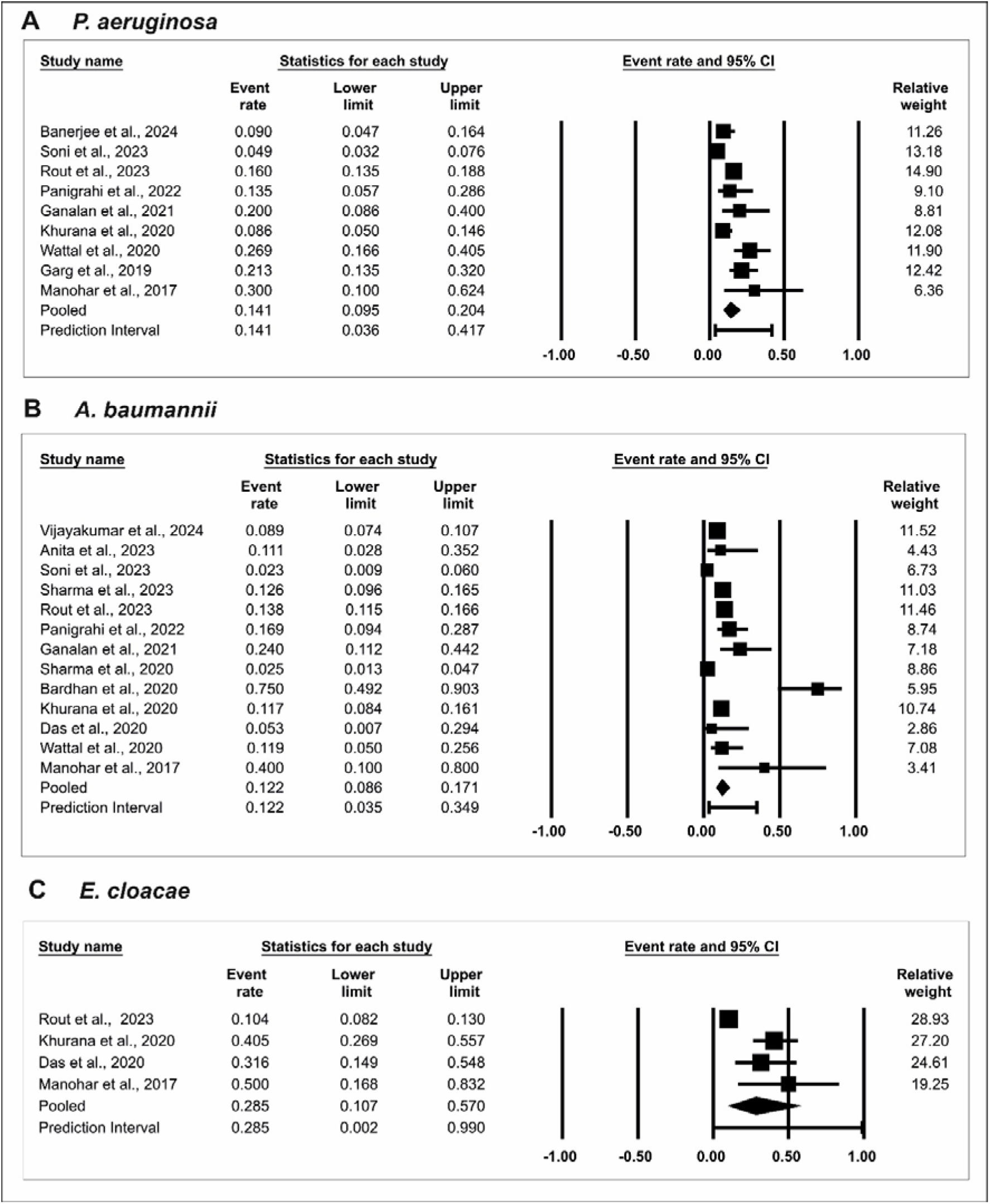
Forest plots demonstrating the pooled prevalence of Col^R^ (A) *P. aeruginosa*, (B) *A. baumannii*, and (C) *E. cloacae*.

Among the major nosocomial pathogens, *K. pneumoniae* showed a very high rate of Col^R^ at 17.4% (95% CI: 11.1 to 26.1) (Figure 2B), followed by *P. aeruginosa* at 14.1% (95% CI: 9.5 to 20.4) (Figure 3A). *E. coli* and *A. baumannii* showed a similar Col^R^ pattern with a resistance rate of 12.3% (95% CI: 6.8 to 21.2) and 12.2 % (95% CI: 8.6 to 17.1) respectively (Figure 2A and 3B). Surprisingly, our study revealed that the bacterial pathogen *E. cloacae* has the highest Col^R^ prevalence rate at 28.5% (95% CI: 10.7 to 50.0) (Figure 3C). However, considering the limited number of studies available and the small number of isolates analysed for *E. cloacae*, further investigation is necessary to get a more robust and accurate estimation of its prevalence in India.

### 3.4 Sensitivity analysis

The pooled rate of Col^R^ among different GNB species was not affected by leave 1-out sensitivity analyses for the bacterial species *E. coli*, *K. pneumoniae*, *P. aeruginosa*, and *A baumannii* (Supplementary Figures 2 and 3). However, for *E.* cloacae, the sensitivity analysis revealed that the study by Rout et al., [20] is significantly influencing the overall Col^R^ rate (Supplementary Figure 3C). Excluding this study would result in a 10% increase in the estimated prevalence. This highlights the importance of considering the robustness of the meta-analysis and the potential influence of individual studies on the overall findings. Further investigation or a more robust analysis strategy might be necessary to strengthen the conclusions.

### 3.5 Geographic distribution of the Col^R^ GNB species in India

On analysing the geographical distribution of the Col^R^ nosocomial GNB species in the Indian subcontinent, it was revealed that the strains were largely reported from the most populated states in India (Figure 4). For example, Col^R^ *K. pneumonia* strains were reported from 11 states and Union Territories, including Rajasthan, Haryana, Delhi, Uttar Pradesh, Bihar, West Bengal, Odisha, Telangana, Tamil Nadu, Kerala, and Chandigarh. Col^R^ *E. coli*, *A. baumannii*, *P. aeruginosa*, and *E. cloacae* were reported from 8, 8, 6, and 4 states respectively. The distribution of different Col^R^ bacterial species in India is shown in Fig. 5. The analysis revealed that Col^R^ is particularly prevalent in the states such as Delhi, Odisha, and Tamil Nadu, as these states harbour all 5 most predominant Col^R^ strains (Figure 4).

**Figure 4.**
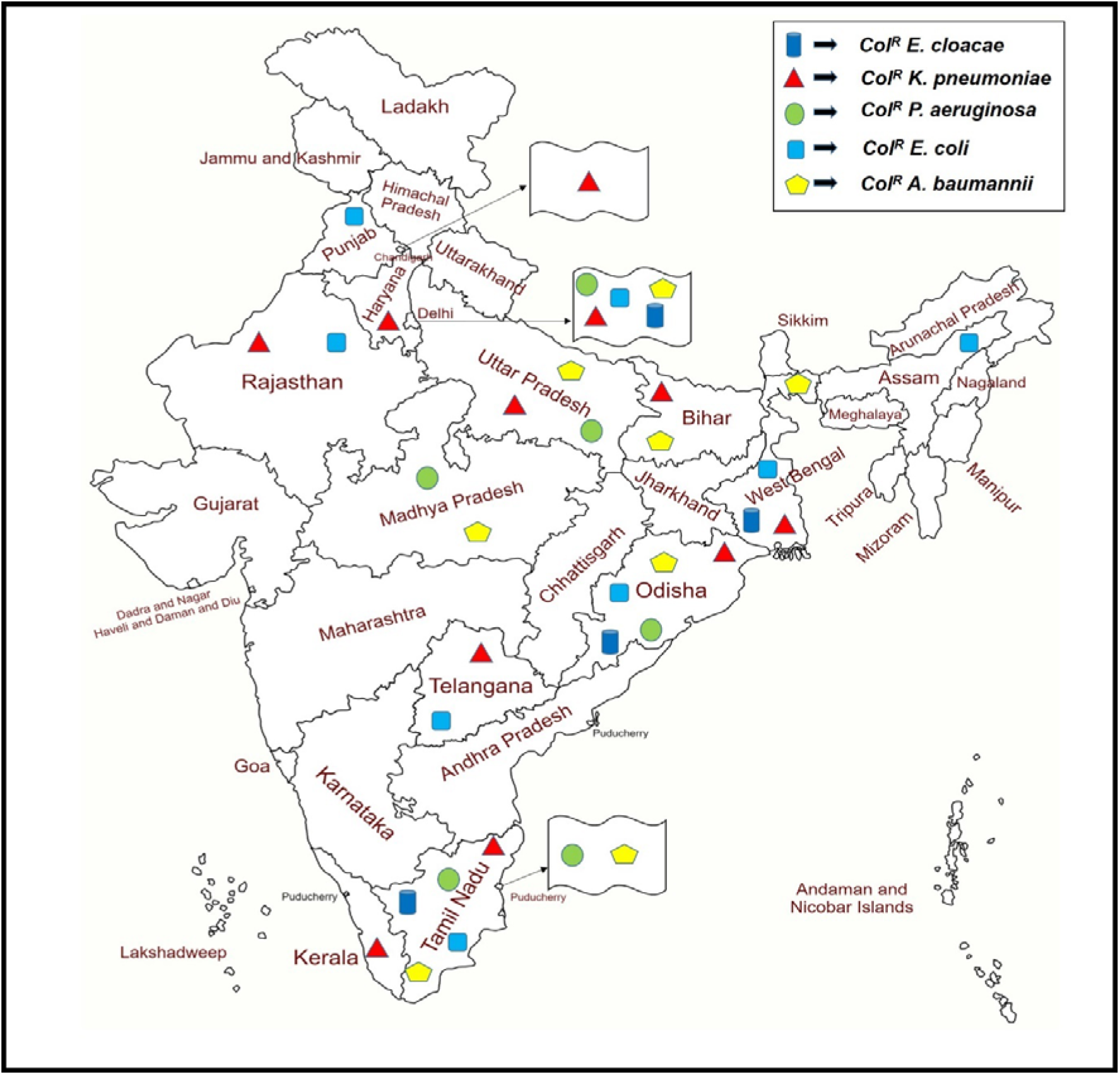
The reports of Col^R^ GNB bacterial species from various states and Union Territories of India.

### 3.6 Col^R^ among the less prevalent GNB species

The study detected that Col^R^ in India is most prevalent among the *E. coli*, *K. pneumoniae*, *P. aeruginosa*, *A. baumannii*, and *E. cloacae* species. Moreover, the study identified Col^R^ in a limited number of other GNB species including *Sphingomonas paucimobilis*, *Raoutella electrica*, *Chryseobacterium gallinarum*, *Citrobacter freundii, Acinetobacter lwoffii, Salmonella enterica, Enterobacter ledwigii*, *Klebsiella oxytoca, Stenotrophomonas maltophilia, Aeromonas verroni, Aeromonas dhakensis, Pseudomonas luteola, Klebsiella quasipneumoniae*, *Hafnia alvei, Enterobacter hormaechei, Aeromonas caviae, Salmonella paratyphi,* and *Enterobacter aerogens* [21,22]. Though the Col^R^ rate among these bacterial species were relatively low, their clinical significance needs to be assessed.

### 3.7 Detection of intrinsic Col^R^ GNB species

Beyond the acquired Col^R^ identified in various studies, India has also reported several bacterial species that are intrinsically resistant to colistin. These GNB species include *Morganella morganii*, *Proteus mirabilis*, *Proteus vulgaris*, *Burkholderia cepacia*, *Serratia marcescens*, *Aeromonas hydrophilia, Burkholderia pseudomalleii*, *Providencia stuaartii, Providencia rettgeri*, and *Vibrio cholerae* [22,23],. Among these intrinsically Col^R^ GNB species, some have been reported to harbour the *mcr* genes, for example in *B. cepacia*, *P. vulgaris*, *P. mirabilis*, *M. morganii,* and *P. rettgeri* [22,24]. As these bacteria can act as a natural reservoir of *mcr* genes, they can potentially facilitate the spread of Col^R^ to other bacterial species through horizontal gene transfer (HGT) mechanisms.

## 4. Discussion

The meta-analysis to determine the level of Col^R^ among the common nosocomial GNB pathogens in India revealed several interesting findings. While *E. cloacae* showed the highest prevalence, there are significantly high levels of Col^R^ in *K. pneumoniae* (17.4%) and *P. aeruginosa* (14.1%) in India (Figures 2 and 3). These bacterial species are notorious for their multi-drug resistance phenotype, the emergence of Col^R^ further complicates the treatment options against them. Recent studies from various countries have reported a low prevalence of Col^R^ in clinical *K. pneumoniae* isolates, notably 1.2% in Ecuador [25], 6.9% in Iran [26], and 4.5% in China [27]. The global prevalence of Col^R^ amongst *K. pneumoniae* isolates from bloodstream infections was reported to be 3.1% [28]. However, few studies have reported a high prevalence of Col^R^ among *P. aeruginosa*, for instance, 21.3% in Egypt [29], and 55% in Taiwan [30].

*E. coli* and *A. baumannii* show moderate levels of Col^R^ in India, which were around 12%. This level of resistance is also significant, especially considering the widespread occurrence of *E. coli* and the potential for *A. baumannii* to cause severe hospital-acquired infections. In *E. coli*, the Col^R^ levels are often very low as reported at 0.5% in Ecuador from faecal samples of humans, pigs, and chickens [25], although higher levels have been observed in Asian countries [31,32]. A recent meta-analysis reported the prevalence of Col^R^ *E. coli* in food samples and food-producing animals to be 5.70% [33]. In South Asian countries such as India, Nepal, Bangladesh, and Pakistan, the pooled prevalence of Col^R^ *E. coli* from poultry was found to be 28% [32]. Another study in China reported the prevalence of Col^R^ *E. coli* during 2016-18 to be around 40% in faecal samples of dogs, cattle, pigs, and chickens [31]. In 2021, a study from Italy reported resistance to colistin in *E. coli* isolated from wild boar to be at 27.9% [34]. The prevalence of Col^R^ *E. coli* in the broiler meat supply chain in Indonesia was found to be 11.76% [35]. A 2022 systematic review found a significant geographic variation in Col^R^ *E. coli* prevalence in clinical samples, with Asia having the highest rate (3.64%), followed by Africa (1.25%), Europe (0.62%), and America (0.48%) [36]. Studies revealed a concerning rise in Col^R^ *E. coli*, particularly in South Asia and China, with animal sources like poultry and livestock showing significantly higher prevalence compared to humans in clinical samples [31,32]. This is significant considering a potential pathway for the spread of Col^R^ *E. coli* from animals to humans through the food chain is possible.

Studies reveal wide variations in Col^R^ prevalence among *A. baumannii* isolates across different regions. Clinical isolates from Egypt report a very high rate at 49% [37], whereas Saudi Arabia showed 8.5% prevalence [38]. Contrastingly, countries like Serbia (3.94%) [39] and Iran (2.8%) [40,41] have shown lower prevalence of Col^R^ *A. baumannii*. A 2024 meta-analysis using the broth microdilution (BMD) method determined a global pooled prevalence of Col^R^ in *A. baumannii* at 4% [42]. It’s important to note that due to *A. baumannii* being a major hospital-acquired pathogen, maximum reports on its Col^R^ have come from clinical settings.

This meta-analysis revealed that among the studied GNB species, *E. cloacae* exhibited the highest prevalence of Col^R^ at 28.5% in India (Figure 3). However, due to the availability of a limited number of reports and isolates of Col^R^ *E. cloacae*, it is difficult to draw definitive conclusions about its prevalence. Therefore, further investigation with a larger dataset is crucial to confirm the extent of Col^R^ in *E. cloacae* in India. While reports on Col^R^ *E. cloacae* are scarce, existing studies reveal significant variations. Clinical isolates in Spain show a prevalence of 4.2% [43], while a report from South Korea shows a much higher rate at 23.9% [44]. Interestingly, a study from Brazil determined a staggering 47% of *E. cloacae* isolates from fish and shrimp that were Col^R^ [45]. This finding suggests a potential reservoir for Col^R^ beyond clinical settings.

As far as mechanisms of Col^R^ development are concerned, primarily the loss-of-function mutations in the genes encoding bacterial TCS systems and their regulons involved in outer membrane remodelling are implicated [1]. Moreover, the spread of plasmid-borne *mcr* genes has a significant role in the rapid emergence of Col^R^ [9]. Most of the studies reported from India have not delved deeply into the mechanistic details of the Col^R^ phenotype. Nevertheless, deleterious mutation in *phoP*, *phoQ*, *pmrA*, *pmrB*, and *mgrB* genes [12,13], complete deletion of *mgrB* [46], presence of efflux pump AcrAB and spermidine export protein MdtI/KpnF and *mcr-1* [47] in *K. pneumoniae*; the presence of *mcr-1* in *E. coli* [48]; mutations at novel sites in *pmrA*, *pmrB*, *lpxA*, *lpxD* genes in *A. baumannii* [49] were reported to be involved in the development of Col^R^ in India. Therefore, more investigations are warranted to understand the molecular details of the reasons behind ColR development.

While the intrinsic resistance to colistin is not a direct risk, the concern lies in their potential to acquire the *mcr* genes through HGT mechanisms. HGT mechanisms allow them to share Col^R^ determinants with other bacteria, potentially rendering them untreatable with the last-resort antibiotic.

## 5. Conclusion

In conclusion, this study highlights the significant rise of Col^R^ in various nosocomial GNB pathogens in India. The level of resistance displayed by *K. pneumoniae*, *P. aeruginosa*, and to a lesser extent, *E. coli* and *A. baumannii*, poses a significant challenge to public health. Though prevalence may differ among regions and over time, continued surveillance, and efforts to curb the spread of resistance are crucial to ensure the continued effectiveness of this critical antibiotic.

## 6. Limitations

This meta-analysis has certain limitations. This study only considered articles published in English, therefore relevant reports in other languages may have been excluded. While the search included major databases like PubMed, Google Scholar, Scopus, and Science Direct, publications indexed in other databases might have been missed. The analysis was restricted to Col^R^ isolates identified employing the BMD method considered to be the gold standard. This potentially excludes resistant strains detected through other, non-BMD methods. Lastly, the present analysis focused largely on the most ubiquitous nosocomial GNB species having Col^R^ phenotype. Therefore, it provides no data on resistance rates among other bacterial types.

## Data Availability

All data produced in the present study are available upon reasonable request to the authors.

## Author Contributions

Conceptualization: SKD, IP, AKP, and SSM; Data extraction and analysis: SKD, IP, AKP, and SSM; Data validation: SKD and IP; Manuscript writing and reviewing: SKD, AKP, and SSM. All authors agreed to the submitted version.

## Acknowledgements

This work is supported by the funds received from the Science and Technology Department, Govt. of Odisha (Grant no. ST-BT-MISC-0005-2023-2463/ST, dt. 23-05-2023). Infrastructure support from the “Centre of Excellence on Bioprospecting of Ethno-pharmaceuticals of Southern Odisha (CoE-BESO)” to the Dept. of Biotechnology, Berhampur University is gratefully acknowledged. Indira Padhy is a recipient of the “Biju Patnaik Research Fellowship (BPRF)” from the Science and Technology Department, Govt. of Odisha.

## References

[1] Padhy I, Dwibedy SK, Mohapatra SS. A molecular overview of the polymyxin-LPS interaction in the context of its mode of action and resistance development. Microbiol Res 2024:127679. 10.1016/j.micres.2024.127679.

[2] Nang SC, Azad MAK, Velkov T, Zhou Q (Tony), Li J. Rescuing the Last-Line Polymyxins: Achievements and Challenges. Pharmacol Rev 2021;73:679–728. 10.1124/pharmrev.120.000020.

[3] Narayan KG, Sinha DK, Singh DK. Escherichia coli. Vet. Public Health Epidemiol., Springer, Singapore; 2023, p. 275–81. 10.1007/978-981-19-7800-5_29.

[4] Bengoechea JA, Sa Pessoa J. *Klebsiella pneumoniae* infection biology: living to counteract host defences. FEMS Microbiol Rev 2019;43:123–44. 10.1093/femsre/fuy043.

[5] Moradali MF, Ghods S, Rehm BHA. Pseudomonas aeruginosa Lifestyle: A Paradigm for Adaptation, Survival, and Persistence. Front Cell Infect Microbiol 2017;7. 10.3389/fcimb.2017.00039.

[6] Morris FC, Dexter C, Kostoulias X, Uddin MI, Peleg AY. The Mechanisms of Disease Caused by Acinetobacter baumannii. Front Microbiol 2019;10:1601. 10.3389/fmicb.2019.01601.

[7] Bannore N, Kapadia F, Hegde A. Difficult to Treat Gram-Negative Bacteria—The Indian Scenario. Curr Infect Dis Rep 2024:1–9. 10.1007/s11908-024-00834-y.

[8] Liu Y-Y, Wang Y, Walsh TR, Yi L-X, Zhang R, Spencer J, et al. Emergence of plasmid-mediated colistin resistance mechanism MCR-1 in animals and human beings in China: a microbiological and molecular biological study. Lancet Infect Dis 2016;16:161–8. 10.1016/S1473-3099(15)00424-7.

[9] Liu J-H, Liu Y-Y, Shen Y-B, Yang J, Walsh TR, Wang Y, et al. Plasmid-mediated colistin-resistance genes: mcr. Trends Microbiol 2024;32:365–78. 10.1016/j.tim.2023.10.006.

[10] Dwibedy SK, Padhy I, Panda AK, Mohapatra SS. Prevalence of polymyxin resistant bacterial strains in India: a systematic review and meta-analysis. J Antimicrob Chemother 2024. 10.1093/jac/dkae130.

[11] Vijayakumar S, Swetha RG, Bakthavatchalam YD, Vasudevan K, Abirami Shankar B, Kirubananthan A, et al. Genomic investigation unveils colistin resistance mechanism in carbapenem-resistant *Acinetobacter baumannii* clinical isolates. Microbiol Spectr 2024;12:e02511–23. 10.1128/spectrum.02511-23.

[12] Nirwan PK, Chatterjee N, Panwar R, Dudeja M, Jaggi N. Mutations in two component system (PhoPQ and PmrAB) in colistin resistant Klebsiella pneumoniae from North Indian tertiary care hospital. J Antibiot (Tokyo) 2021;74:450–7. 10.1038/s41429-021-00417-2.

[13] Mathur P, Khurana S, De Man TJB, Rastogi N, Katoch O, Veeraraghavan B, et al. Multiple importations and transmission of colistin-resistant *Klebsiella pneumoniae* in a hospital in northern India. Infect Control Hosp Epidemiol 2019;40:1387–93. 10.1017/ice.2019.252.

[14] Page MJ, McKenzie JE, Bossuyt PM, Boutron I, Hoffmann TC, Mulrow CD, et al. The PRISMA 2020 statement: an updated guideline for reporting systematic reviews. Syst Rev 2021;10:89. 10.1186/s13643-021-01626-4.

[15] Panda AK. Maintaining ethics, Integrity, and accountability: Best practices for reporting a meta-analysis. Account Res 2024;0:1–3. 10.1080/08989621.2024.2334722.

[16] Higgins JPT, Thompson SG, Deeks JJ, Altman DG. Measuring inconsistency in meta-analyses. BMJ 2003;327:557. 10.1136/bmj.327.7414.557.

[17] Higgins JPT, Green S, editors. Cochrane Handbook for Systematic Reviews of Interventions. The Cochrane Collaboration; 2011.

[18] Sterne JAC, Egger M. Funnel plots for detecting bias in meta-analysis: Guidelines on choice of axis. J Clin Epidemiol 2001;54:1046–55. 10.1016/S0895-4356(01)00377-8.

[19] Egger M, Smith GD, Schneider M, Minder C. Bias in meta-analysis detected by a simple, graphical test. BMJ 1997;315:629. 10.1136/bmj.315.7109.629.

[20] Rout B, Dash SK, Sahu KK, Behera B, Praharaj I, Otta S. Evaluation of different methods for in vitro susceptibility testing of colistin in carbapenem resistant Gram negative bacilli. 2023. 10.1099/acmi.0.000595.v1.

[21] Gaur M, Dey S, Sahu A, Dixit S, Sarathbabu S, Zothanzama J, et al. Characterization and Comparative Genomic Analysis of a Highly Colistin-Resistant Chryseobacterium gallinarum: a Rare, Uncommon Pathogen. Front Cell Infect Microbiol 2022;12:933006. 10.3389/fcimb.2022.933006.

[22] Mitra S, Basu S, Rath S, Sahu SK. Colistin resistance in Gram-negative ocular infections: prevalence, clinical outcome and antibiotic susceptibility patterns. Int Ophthalmol 2020;40:1307–17. 10.1007/s10792-020-01298-4.

[23] Ramesh N, Prasanth M, Ramkumar S, Suresh M, Tamhankar AJ. Colistin susceptibility of gram-negative clinical isolates from Tamil Nadu, India 2016;10.

[24] Premnath MAC, Prabakaran K, Sivasankar S, Boppe A, Sriramajayam L, Jeyaraj S. Occurrence of mcr genes and alterations in mgrB gene in intrinsic colistin-resistant Enterobacterales isolated from chicken meat samples. Int J Food Microbiol 2023;404:110323. 10.1016/j.ijfoodmicro.2023.110323.

[25] Bastidas-Caldes C, Guerrero-Freire S, Ortuño-Gutiérrez N, Sunyoto T, Gomes-Dias CA, Ramírez MS, et al. Colistin resistance in Escherichia coli and Klebsiella pneumoniae in humans and backyard animals in Ecuador. Rev Panam Salud Pública 2023;47:1. 10.26633/RPSP.2023.48.

[26] Narimisa N, Goodarzi F, Bavari S. Prevalence of colistin resistance of Klebsiella pneumoniae isolates in Iran: a systematic review and meta-analysis. Ann Clin Microbiol Antimicrob 2022;21:29. 10.1186/s12941-022-00520-8.

[27] Liu Y, Lin Y, Wang Z, Hu N, Liu Q, Zhou W, et al. Molecular Mechanisms of Colistin Resistance in Klebsiella pneumoniae in a Tertiary Care Teaching Hospital. Front Cell Infect Microbiol 2021;11:673503. 10.3389/fcimb.2021.673503.

[28] Uzairue LI, Rabaan AA, Adewumi FA, Okolie OJ, Folorunso JB, Bakhrebah MA, et al. Global Prevalence of Colistin Resistance in Klebsiella pneumoniae from Bloodstream Infection: A Systematic Review and Meta-Analysis. Pathogens 2022;11:1092. 10.3390/pathogens11101092.

[29] El-Baky RMA, Masoud SM, Mohamed DS, Waly NG, Shafik EA, Mohareb DA, et al. Prevalence and Some Possible Mechanisms of Colistin Resistance Among Multidrug-Resistant and Extensively Drug-Resistant Pseudomonas aeruginosa. Infect Drug Resist 2020;13:323. 10.2147/IDR.S238811.

[30] Liu P-Y, Weng L-L, Tseng S-Y, Huang C-C, Cheng C-C, Mao Y-C, et al. Colistin Resistance of *Pseudomonas aeruginosa* Isolated from Snakes in Taiwan. Can J Infect Dis Med Microbiol 2017;2017:1–5. 10.1155/2017/7058396.

[31] Li F, Cheng P, Li X, Liu R, Liu H, Zhang X. Molecular Epidemiology and Colistin-Resistant Mechanism of mcr-Positive and mcr-Negative Escherichia coli Isolated From Animal in Sichuan Province, China. Front Microbiol 2022;13:818548. 10.3389/fmicb.2022.818548.

[32] Dawadi P, Bista S, Bista S. Prevalence of Colistin-Resistant Escherichia coli from Poultry in South Asian Developing Countries. Vet Med Int 2021;2021:1–5. 10.1155/2021/6398838.

[33] Lencina FA, Bertona M, Stegmayer MA, Olivero CR, Frizzo LS, Zimmermann JA, et al. Prevalence of colistin-resistant Escherichia coli in foods and food-producing animals through the food chain: A worldwide systematic review and meta-analysis. Heliyon 2024;10:e26579. 10.1016/j.heliyon.2024.e26579.

[34] Cilia G, Turchi B, Fratini F, Ebani VV, Turini L, Cerri D, et al. Phenotypic and genotypic resistance to colistin in E. coli isolated from wild boar (Sus scrofa) hunted in Italy. Eur J Wildl Res 2021;67:57. 10.1007/s10344-021-01501-6.

[35] Palupi MF, Wibawan IWT, Sudarnika E, Maheshwari H, Darusman HS. PREVALENCE OF mcr-1 COLISTIN RESISTANT GENE IN Escherichia coli ALONG THE BROILER MEAT SUPPLY CHAIN IN INDONESIA. BIOTROPIA 2019;26:143–53. 10.11598/btb.2019.26.2.1054.

[36] Dadashi M, Sameni F, Bostanshirin N, Yaslianifard S, Khosravi-Dehaghi N, Nasiri MJ, et al. Global prevalence and molecular epidemiology of mcr-mediated colistin resistance in Escherichia coli clinical isolates: a systematic review. J Glob Antimicrob Resist 2022;29:444–61. 10.1016/j.jgar.2021.10.022.

[37] Seleim SM, Mostafa MS, Ouda NH, Shash RY. The role of pmrCAB genes in colistin-resistant Acinetobacter baumannii. Sci Rep 2022;12:20951. 10.1038/s41598-022-25226-x.

[38] Elham B, Fawzia A. Colistin resistance in Acinetobacter baumannii isolated from critically ill patients: clinical characteristics, antimicrobial susceptibility and outcome. Afr Health Sci 2019;19:2400–6. 10.4314/ahs.v19i3.13.

[39] Kabic J, Novovic K, Kekic D, Trudic A, Opavski N, Dimkic I, et al. Comparative genomics and molecular epidemiology of colistin-resistant Acinetobacter baumannii. Comput Struct Biotechnol J 2023;21:574–85. 10.1016/j.csbj.2022.12.045.

[40] Khoshbayan A, Shariati A, Shahmoradi S, Baseri Z, Mozafari H, Darban-Sarokhalil D. Prevalence and molecular mechanisms of colistin resistance in Acinetobacter baumannii clinical isolates in Tehran, Iran. Acta Microbiol Immunol Hung 2021. 10.1556/030.2021.01420.

[41] Khoshnood S, Savari M, Abbasi Montazeri E, Farajzadeh Sheikh A. Survey on Genetic Diversity, Biofilm Formation, and Detection of Colistin Resistance Genes in Clinical Isolates of Acinetobacter baumannii. Infect Drug Resist 2020;Volume 13:1547–58. 10.2147/IDR.S253440.

[42] Bostanghadiri N, Narimisa N, Mirshekar M, Dadgar-Zankbar L, Taki E, Navidifar T, et al. Prevalence of colistin resistance in clinical isolates of Acinetobacter baumannii: a systematic review and meta-analysis. Antimicrob Resist Infect Control 2024;13:24. 10.1186/s13756-024-01376-7.

[43] Prim N, Turbau M, Rivera A, Rodríguez-Navarro J, Coll P, Mirelis B. Prevalence of colistin resistance in clinical isolates of Enterobacteriaceae: A four-year cross-sectional study. J Infect 2017;75:493–8. 10.1016/j.jinf.2017.09.008.

[44] Hong Y-K, Lee J-Y, Ko KS. Colistin resistance in Enterobacter spp. isolates in Korea. J Microbiol 2018;56:435–40. 10.1007/s12275-018-7449-0.

[45] Almeida MVAD, Brito ILP, Carvalho ALSD, Costa RA. In vitro resistance of Enterobacter cloacae isolated from fresh seafood to colistin. Rev Soc Bras Med Trop 2018;51:674–5. 10.1590/0037-8682-0287-2017.

[46] Bandyopadhyay S, Bhattacharyya D, Samanta I, Banerjee J, Habib M, Dutta TK, et al. Characterization of Multidrug-Resistant Biofilm-Producing *Escherichia coli* and *Klebsiella pneumoniae* in Healthy Cattle and Cattle with Diarrhea. Microb Drug Resist 2021;27:1457–69. 10.1089/mdr.2020.0298.

[47] Bir R, Gautam H, Arif N, Chakravarti P, Verma J, Banerjee S, et al. Analysis of colistin resistance in carbapenem-resistant *Enterobacterales* and XDR *Klebsiella pneumoniae*. Ther Adv Infect Dis 2022;9:204993612210806. 10.1177/20499361221080650.

[48] Elizabeth R, Baishya S, Kalita B, Wangkheimayum J, Choudhury MD, Chanda DD, et al. Colistin exposure enhances expression of eptB in colistin-resistant Escherichia coli co-harboring mcr-1. Sci Rep 2022;12:1348. 10.1038/s41598-022-05435-0.

[49] Sharma S, Banerjee T, Yadav G, Palandurkar K. Mutations at Novel Sites in *pmrA/B* and *lpxA/D* Genes and Absence of Reduced Fitness in Colistin-Resistant *Acinetobacter baumannii* from a Tertiary Care Hospital, India. Microb Drug Resist 2021;27:628–36. 10.1089/mdr.2020.0023.

