## Supplementary Information for "Colistin resistance among the Gram-negative nosocomial pathogens in India: A systematic review and meta-analysis"

**Running Title:** Colistin-resistant nosocomial pathogens in India

**Sambit K. Dwibedy**[1,2], **Indira Padhy**[1], **Aditya K. Panda**[1,3], **Saswat S. Mohapatra**[1,3]*

[1]Department of Biotechnology, Berhampur University, Bhanja Bihar, Berhampur- 760007, Odisha, India

[2]Department of Zoology, SBRG Women's College, Berhampur- 760001, Odisha, India

[3]Centre of Excellence on Bioprospecting of Ethno-pharmaceuticals of Southern Odisha (CoE-BESO), Berhampur University, Bhanja Bihar, Berhampur- 760007, India

*** Corresponding author**

**Saswat S. Mohapatra**, PhD
Assistant Professor, Department of Biotechnology, Berhampur University, Bhanja Bihar, Berhampur- 760007, India.

**Supplementary Table 1.** Reports of $Col^R$ *K. pneumoniae* from India with source, sample size, colistin resistant isolate and *mcr* involvement.

| **Sl. No.** | **Author and Year** | **State / UT** | **Source** | **Sample size** | **No. of $Col^R$ isolates** | ***mcr*** |
|---|---|---|---|---|---|---|
| 1 | Vasesi et al., 2024 [1] | Chandigarh | Clinical | 50 | 3 | - |
| 2 | Borde et al., 2023[2] | Telangana | Clinical | 103 | 11 | - |
| 3 | Anita et al., 2023 [3] | Bihar | Clinical | 48 | 1 | - |
| 4 | Kaza et al., 2023 [4] | Chandigarh | Clinical | 775 | 18 | - |
| 5 | Ranjan et al., 2023 [5] | Telangana | Clinical | 66 | 6 | - |
| 6 | Shanthini et al., 2023 [6] | Tamil Nadu | Clinical | 30 | 27 | - |
| 7 | Rout et al., 2023 [7] | Odisha | Clinical | 1690 | 319 | - |
| 8 | Sharma et al., 2022 [8] | Uttar Pradesh | Clinical | 45 | 31 | - |
| 9 | Bir et al., 2022 [9] | Delhi | Clinical | 48 | 7 | *mcr-1* +ve |
| 10 | Panigrahi et al., 2022 [10] | Odisha | Clinical | 199 | 33 | - |
| 11 | Das et al., 2021 [11] | Odisha, UP, Rajasthan | Clinical | 158 | 27 | - |
| 12 | Nirwan et al., 2021 [12] | Haryana | Clinical | 1689 | 13 | - |
| 13 | Kar et al., 2021 [13] | Odisha | Clinical | 79 | 22 | - |
| 14 | Bandyopadhyay et al., 2021 [14] | West Bengal | Diary | 37 | 1 | - |
| 15 | Bardhan et al., 2020 [15] | West Bengal | Environmental | 18 | 15 | - |
| 16 | Khurana et al., 2020 [16] | Delhi | Clinical | 245 | 102 | - |
| 17 | Das et al., 2020 [17] | West Bengal | Clinical | 56 | 22 | - |
| 18 | Wattal et al., 2020 [18] | Delhi | Clinical | 109 | 52 | - |
| 19 | Mathur et al., 2019 [19] | Delhi | Clinical | 846 | 34 | - |
| 20 | Sundaramoorthy et al., 2019 A [20] | Tamil Nadu | Clinical | 6 | 3 | - |
| 21 | Raghupati et al., 2019 [21] | Tamil Nadu | Clinical | 48 | 19 | - |
| 22 | Behera et al., 2018 [22] | Odisha | Clinical | 55 | 4 | - |
| 23 | Kumar et al., 2018 [23] | Kerala | Clinical | 932 | 17 | - |
| 24 | Manohar et al., 2017 [24] | Tamil Nadu | Clinical | 18 | 6 | - |
| Total | | | | 7350 | 793 | - |

**Supplementary Table 2**. Reports of Col[R] *E. cloacae* from India with source, sample size, colistin resistant isolate and *mcr* involvement.

| **Sl. No.** | **Author and Year** | **State** | **Source** | **Sample Size** | **No of Col[R] Isolate** | ***mcr*** |
|---|---|---|---|---|---|---|
| 1 | Rout et al., 2023 [7] | Odisha | Clinical | 635 | 66 | - |
| 2 | Khurana et al., 2020 [16] | Delhi | Clinical | 42 | 17 | - |
| 3 | Das et al., 2020 [17] | West Bengal | Clinical | 19 | 6 | - |
| 4 | Manohar et al., 2017 [24] | Tamil Nadu | Clinical | 6 | 3 | - |
| Total | | | | 702 | 92 | - |

**Supplementary Table 3.** Reports of Col[R] *P. aeruginosa* from India with source, sample size, colistin resistant isolate and *mcr* involvement.

| **Sl. No.** | **Author and Year** | **State** | **Source** | **Sample Size** | **No of Col[R] Isolate** | ***mcr*** |
|---|---|---|---|---|---|---|
| 1 | Banerjee et al., 2024 [25] | Uttar Pradesh | Clinical | 100 | 9 | - |
| 2 | Soni et al., 2023 [26] | Madhya Pradesh | Clinical | 384 | 19 | - |
| 3 | Rout et al., 2023 [7] | Odisha | Clinical | 731 | 117 | - |
| 4 | Panigrahi et al., 2022 [10] | Odisha | Clinical | 37 | 5 | - |
| 5 | Gunalan et al., 2021 [27] | Pondicherry | Clinical | 25 | 5 | - |
| 6 | Khurana et al., 2020 [16] | Delhi | Clinical | 139 | 12 | - |
| 7 | Wattal et al., 2020 [18] | Delhi | Clinical | 52 | 14 | - |
| 8 | Garg et al., 2019 [28] | UP | Clinical | 75 | 16 | - |
| 9 | Manohar et al., 2017 [24] | Tamil Nadu | Clinical | 10 | 3 | - |
| Total | | | | 1553 | 200 | - |

**Supplementary Table 4.** Reports of $Col^R$ *E. coli* from India with source, sample size, colistin resistant isolate and *mcr* involvement.

| Sl. No. | Author and Year | State | Source | Sample Size | No of $Col^R$ Isolate | *mcr* |
|---|---|---|---|---|---|---|
| 1 | Satapathy et al., 2023 [29] | Punjab | Goat milk | 22 | 11 | - |
| 2 | Ranjan et al., 2023 [5] | Telangana | Clinical | 28 | 1 | - |
| 3 | Rout et al., 2023 [7] | Odisha | Clinical | 2255 | 179 | - |
| 4 | Elizabeth et al., 2022 [30] | Assam | Clinical | 291 | 12 | *mcr-1*(5) |
| 5 | Bir et al., 2022 [9] | Delhi | Clinical | 50 | 8 | - |
| 6 | Panigrahi et al., 2022 [10] | Odisha | Clinical | 107 | 18 | - |
| 7 | Kar et al., 2021 [13] | Odisha | Clinical | 113 | 3 | - |
| 8 | Priyanka et al., 2021 [31] | Rajasthan | Leafy green | 117 | 62 | - |
| 9 | Bandyopadhyay et al., 2021 [14] | West Bengal | Diary | 35 | 1 | - |
| 10 | Bardhan et al., 2020 [15] | West Bengal | Environmental | 20 | 14 | - |
| 11 | Khurana et al., 2020 [16] | Delhi | Clinical | 158 | 8 | - |
| 12 | Das et al., 2020 [17] | West Bengal | Clinical | 27 | 2 | - |
| 13 | Behera et al., 2018 [22] | Odisha | Clinical | 99 | 3 | - |
| 14 | Wattal et al., 2020 [18] | Delhi | Clinical | 22 | 2 | - |
| 15 | Sundaramoorthy et al., 2019 B [32] | Tamil Nadu | Clinical | 8 | 2 | - |
| 16 | Amladi et al., 2019 [33] | Tamil Nadu | Clinical | 81 | 4 | - |
| 17 | Manohar et al., 2017 [24] | Tamil Nadu | Clinical | 43 | 11 | - |
| Total | | | | 3476 | 341 | - |

**Supplementary Table 5.** Reports of $Col^R$ *A. baumannii* from India with source, sample size, colistin resistant isolate and *mcr* involvement.

| **Sl. No.** | **Author and Year** | **State** | **Source** | **Sample Size** | **No of $Col^R$ Isolate** | ***mcr*** |
|---|---|---|---|---|---|---|
| 1 | Vijayakumar et al., 2024 [34] | Tamil Nadu | Clinical | 1212 | 108 | - |
| 2 | Soni et al., 2023 [26] | Madhya Pradesh | Clinical | 174 | 4 | - |
| 3 | Anita et al., 2023 [3] | Bihar | Clinical | 18 | 2 | - |
| 4 | Sharma et al., 2023 [35] | Uttar Pradesh | Clinical | 356 | 45 | - |
| 5 | Rout et al., 2023 [7] | Odisha | Clinical | 702 | 97 | - |
| 6 | Panigrahi et al., 2022 [10] | Odisha | Clinical | 59 | 10 | - |
| 7 | Gunalan et al., 2021 [27] | Pondicherry | Clinical | 25 | 6 | - |
| 8 | Sharma et al., 2021 [36] | Uttar Pradesh | Clinical | 365 | 9 | - |
| 9 | Bardhan et al., 2020 [15] | West Bengal | Environmental | 16 | 12 | - |
| 10 | Khurana et al., 2020 [16] | Delhi | Clinical | 273 | 32 | - |
| 11 | Das et al., 2020 [17] | West Bengal | Clinical | 19 | 1 | - |
| 12 | Wattal et al., 2020 [18] | Delhi | Clinical | 42 | 5 | - |
| 13 | Manohar et al., 2017 [24] | Tamil Nadu | Clinical | 5 | 2 | |
| Total | | | | 3266 | 333 | - |

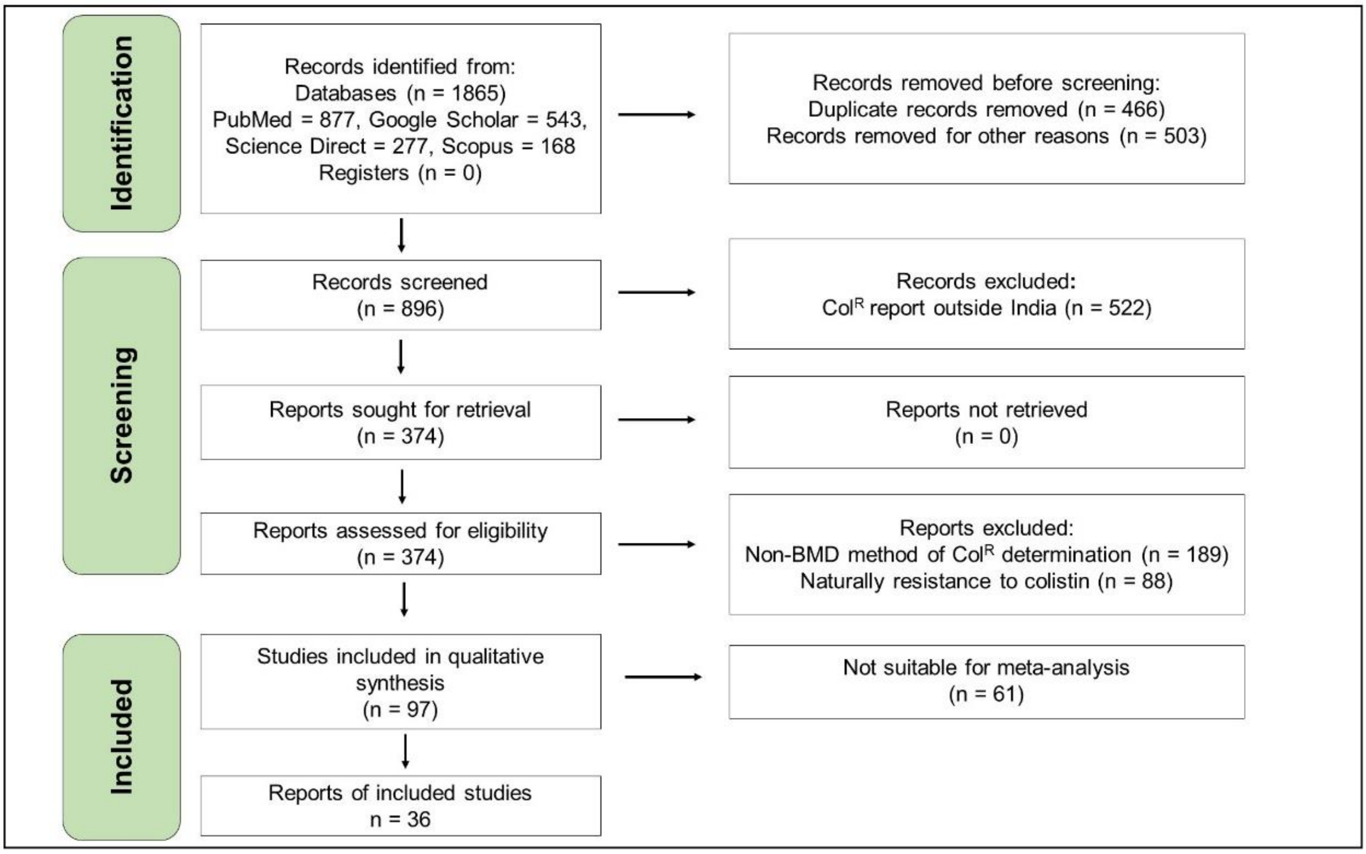


**Supplementary Figure 1.** The PRISMA flow diagram for the selection, inclusion, and exclusion of studies.

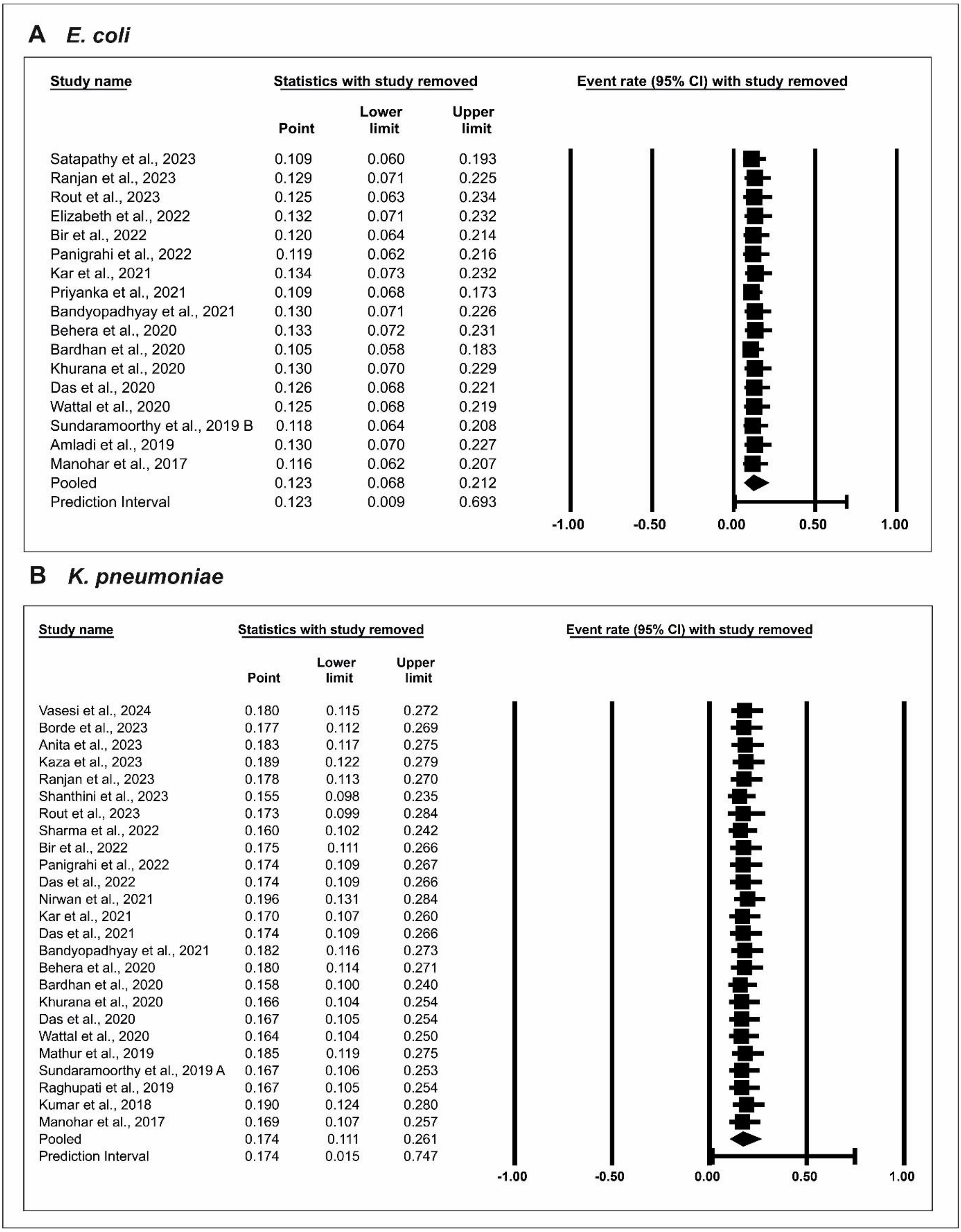


**Supplementary Figure 2**. Sensitivity plots for (A) *E. coli*, and (B) *K. pneumoniae*.

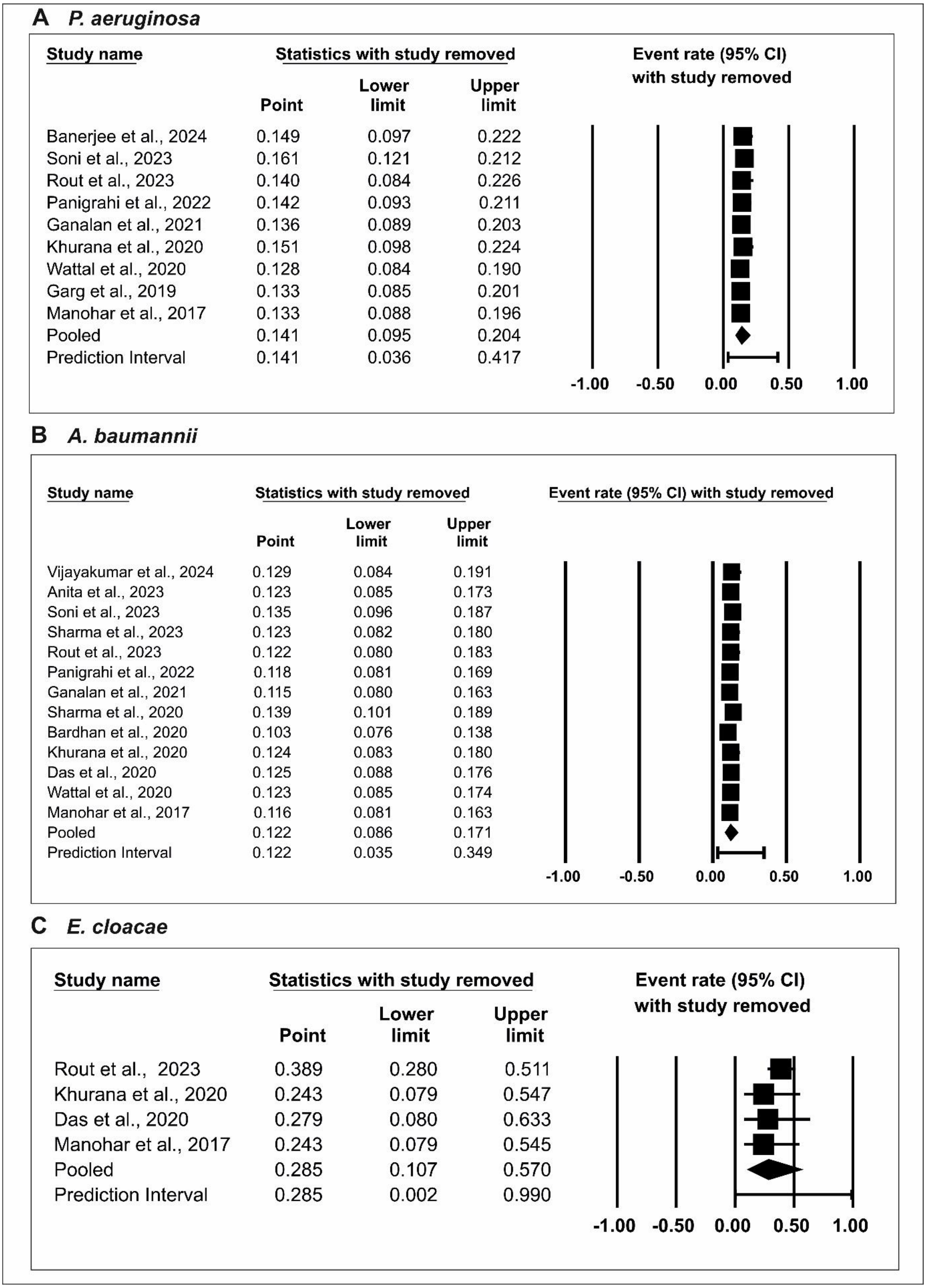


**Supplementary Figure 3**. Sensitivity plots for (A) *P. aeruginosa*, (B) *A. baumannii*, and (C) *E. cloacae* in India.

### References


[1] Vasesi D, Gupta V, Gupta P, Singhal L. Risk factor and resistance profile of colistin resistant Acinetobacter baumannii and Klebsiella pneumoniae. Indian J Med Microbiol 2024;47:100486. https://doi.org/10.1016/j.ijmmb.2023.100486.

[2] Borde K, Kareem MA, Sharma RM, Dass SM, Ravi V, Mathai D. *In vitro* activity of cefiderocol against comparators (ceftazidime-avibactam, ceftazidime-avibactam/ aztreonam combination, and colistin) against clinical isolates of meropenem-resistant *Klebsiella pneumoniae* from India. Microbiol Spectr 2023;11:e00847-23. https://doi.org/10.1128/spectrum.00847-23.

[3] . A, Kumari R, Saurabh K, Kumar S, Kumari N. Comparative Evaluation of Broth Microdilution With Disc Diffusion and VITEK 2 for Susceptibility Testing of Colistin on Multidrug-Resistant Gram-Negative Bacteria. Cureus 2023. https://doi.org/10.7759/cureus.50894.

[4] Kaza P, Xavier BB, Mahindroo J, Singh N, Baker S, Nguyen TNT, et al. Extensively Drug-Resistant *Klebsiella pneumoniae* Associated with Complicated Urinary Tract Infection in Northern India. Jpn J Infect Dis 2024;77:7–15. https://doi.org/10.7883/yoken.JJID.2023.009.

[5] Ranjan R, Iyer RN, Jangam RR, Arora N. Evaluation of in-vitro colistin susceptibility and clinical profile of carbapenem resistant Enterobacteriaceae related invasive infections. Indian J Med Microbiol 2023;41:40–4. https://doi.org/10.1016/j.ijmmb.2022.12.008.

[6] Shanthini T, Manohar P, Hua X, Leptihn S, Nachimuthu R. Detection of Hypervirulent *Klebsiella pneumoniae* from Clinical Samples in Tamil Nadu. Infectious Diseases (except HIV/AIDS); 2023. https://doi.org/10.1101/2023.02.19.23286158.

[7] Rout B, Dash SK, Sahu KK, Behera B, Praharaj I, Otta S. Evaluation of different methods for in vitro susceptibility testing of colistin in carbapenem resistant Gram negative bacilli. 2023. https://doi.org/10.1099/acmi.0.000595.v1.

[8] Sharma S, Banerjee T, Kumar A, Yadav G, Basu S. Extensive outbreak of colistin resistant, carbapenemase (blaOXA-48, blaNDM) producing Klebsiella pneumoniae in a large tertiary care hospital, India. Antimicrob Resist Infect Control 2022;11:1. https://doi.org/10.1186/s13756-021-01048-w.

[9] Bir R, Gautam H, Arif N, Chakravarti P, Verma J, Banerjee S, et al. Analysis of colistin resistance in carbapenem-resistant *Enterobacterales* and XDR *Klebsiella pneumoniae*. Ther Adv Infect Dis 2022;9:204993612210806. https://doi.org/10.1177/20499361221080650.

[10] Panigrahi K, Pathi BK, Poddar N, Sabat S, Pradhan S, Pattnaik D, et al. Colistin Resistance Among Multi-Drug Resistant Gram-Negative Bacterial Isolates From Different Clinical Samples of ICU Patients: Prevalence and Clinical Outcomes. Cureus 2022. https://doi.org/10.7759/cureus.28317.

[11] Das A, Sahoo RK, Gaur M, Dey S, Sahoo S, Sahu A, et al. Molecular prevalence of resistance determinants, virulence factors and capsular serotypes among colistin resistance carbapenemase producing Klebsiella pneumoniae: a multi-centric retrospective study. 3 Biotech 2022;12:30. https://doi.org/10.1007/s13205-021-03056-4.

[12] Nirwan PK, Chatterjee N, Panwar R, Dudeja M, Jaggi N. Mutations in two component system (PhoPQ and PmrAB) in colistin resistant Klebsiella pneumoniae from North

Indian tertiary care hospital. J Antibiot (Tokyo) 2021;74:450–7. https://doi.org/10.1038/s41429-021-00417-2.

[13] Kar P, Behera B, Mohanty S, Jena J, Mahapatra A. Detection of Colistin Resistance in Carbapenem Resistant Enterobacteriaceae by Reference Broth Microdilution and Comparative Evaluation of Three Other Methods. J Lab Physicians 2021;13:263–9. https://doi.org/10.1055/s-0041-1731137.

[14] Bandyopadhyay S, Bhattacharyya D, Samanta I, Banerjee J, Habib M, Dutta TK, et al. Characterization of Multidrug-Resistant Biofilm-Producing *Escherichia coli* and *Klebsiella pneumoniae* in Healthy Cattle and Cattle with Diarrhea. Microb Drug Resist 2021;27:1457–69. https://doi.org/10.1089/mdr.2020.0298.

[15] Bardhan T, Chakraborty M, Bhattacharjee B. Prevalence of Colistin-Resistant, Carbapenem-Hydrolyzing Proteobacteria in Hospital Water Bodies and Out-Falls of West Bengal, India. Int J Environ Res Public Health 2020;17:1007. https://doi.org/10.3390/ijerph17031007.

[16] Khurana S, Malhotra R, Mathur P. Evaluation of Vitek®2 performance for colistin susceptibility testing for Gram-negative isolates. JAC-Antimicrob Resist 2020;2:dlaa101. https://doi.org/10.1093/jacamr/dlaa101.

[17] Das S, Roy S, Roy S, Goelv G, Sinha S, Mathur P, et al. Colistin Susceptibility Testing of Gram-Negative Bacilli: Better Performance of Vitek2 System than E-Test Compared to Broth Microdilution Method as the Gold Standard Test. Indian J Med Microbiol 2020;38:58–65. https://doi.org/10.4103/ijmm.IJMM_19_480.

[18] Wattal C, Goel N, Oberoi JK, Datta S, Raveendran R. Performance of Three Commercial Assays for Colistin Susceptibility in Clinical Isolates and Mcr-1 Carrying Reference Strain. Indian J Med Microbiol 2019;37:488–95. https://doi.org/10.4103/ijmm.IJMM_20_92.

[19] Mathur P, Khurana S, De Man TJB, Rastogi N, Katoch O, Veeraraghavan B, et al. Multiple importations and transmission of colistin-resistant *Klebsiella pneumoniae* in a hospital in northern India. Infect Control Hosp Epidemiol 2019;40:1387–93. https://doi.org/10.1017/ice.2019.252.

[20] Sundaramoorthy NS, Mohan HM, Subramaniam S, Raman T, Selva Ganesan S, Sivasubamanian A, et al. Ursolic acid inhibits colistin efflux and curtails colistin resistant Enterobacteriaceae. AMB Express 2019;9:27. https://doi.org/10.1186/s13568-019-0750-4.

[21] Ragupathi ND, Bakthavatchalam Y, Mathur P, Pragasam A, Walia K, Ohri V, et al. Plasmid profiles among some ESKAPE pathogens in a tertiary care centre in south India. Indian J Med Res 2019;149:222. https://doi.org/10.4103/ijmr.IJMR_2098_17.

[22] Behera B, Jena J, Kar P, Mohanty S, Mahapatra A. Deciphering Polymyxin B Minimum Inhibitory Concentration from Colistin Minimum Inhibitory Concentration and Vice Versa: An Analysis on 156 Carbapenem-Resistant Enterobacteriaceae Isolates. Indian J Med Microbiol 2018;36:587–9. https://doi.org/10.4103/ijmm.IJMM_18_293.

[23] Kumar A, Biswas L, Omgy N, Mohan K, Vinod V, Sajeev A, et al. Colistin resistance due to insertional inactivation of the mgrB in Klebsiella pneumoniae of clinical origin: First report from India: Resistencia a colistina debido a inactivación insercional del gen mgrB en aislados clínicos de Klebsiella pneumoniae: Primera notificación en India. Rev Esp Quimioter 2018;31:406.

[24] Manohar P, Shanthini T, Ayyanar R, Bozdogan B, Wilson A, Tamhankar AJ, et al. The distribution of carbapenem- and colistin-resistance in Gram-negative bacteria from the Tamil Nadu region in India. J Med Microbiol 2017;66:874–83. https://doi.org/10.1099/jmm.0.000508.

[25] Banerjee T, Adwityama A, Sharma S, Mishra K, Prusti P, Maitra U. Comparative evaluation of colistin broth disc elution (CBDE) and broth microdilution (BMD) in clinical isolates of Pseudomonas aeruginosa with special reference to heteroresistance. Indian J Med Microbiol 2024;47:100494. https://doi.org/10.1016/j.ijmmb.2023.100494.

[26] Soni M, Kapoor G, Perumal N, Chaurasia D. Emergence of Multidrug-Resistant Non-Fermenting Gram-Negative Bacilli in a Tertiary Care Teaching Hospital of Central India: Is Colistin Resistance Still a Distant Threat? Cureus 2023. https://doi.org/10.7759/cureus.39243.

[27] Gunalan A, Sarumathi D, Sastry AS, Ramanathan V, Rajaa S, Sistla S. Effect of combined colistin and meropenem against meropenem resistant Acinetobacter baumannii and Pseudomonas aeruginosa by checkerboard method: A cross sectional analytical study n.d.

[28] Garg A, Garg J, Kumar S, Bhattacharya A, Agarwal S, Upadhyay G. Molecular epidemiology & therapeutic options of carbapenem-resistant Gram-negative bacteria. Indian J Med Res 2019;149:285. https://doi.org/10.4103/ijmr.IJMR_36_18.

[29] Satpathy MM, Sharma NS, Kaur P, Arora AK. Detection of antimicrobial resistance genes in extended spectrum beta-lactamase-producing Escherichia coli from milk of indigenous Beetal goats of Punjab. Iran J Vet Res 2023;24. https://doi.org/10.22099/ijvr.2023.43480.6365.

[30] Elizabeth R, Baishya S, Kalita B, Wangkheimayum J, Choudhury MD, Chanda DD, et al. Colistin exposure enhances expression of eptB in colistin-resistant Escherichia coli co-harboring mcr-1. Sci Rep 2022;12:1348. https://doi.org/10.1038/s41598-022-05435-0.

[31] Priyanka, Meena PR, Meghwanshi KK, Rana A, Singh AP. Leafy greens as a potential source of multidrug-resistant diarrhoeagenic Escherichia coli and Salmonella. Microbiology 2021;167. https://doi.org/10.1099/mic.0.001059.

[32] Sundaramoorthy NS, Suresh P, Selva Ganesan S, GaneshPrasad A, Nagarajan S. Restoring colistin sensitivity in colistin-resistant E. coli: Combinatorial use of MarR inhibitor with efflux pump inhibitor. Sci Rep 2019;9:19845. https://doi.org/10.1038/s41598-019-56325-x.

[33] Amladi A, Abirami B, Devi Sm, Sudarsanam T, Kandasamy S, Kekre N, et al. Susceptibility profile, resistance mechanisms & efficacy ratios of fosfomycin, nitrofurantoin & colistin for carbapenem-resistant Enterobacteriaceae causing urinary tract infections. Indian J Med Res 2019;149:185. https://doi.org/10.4103/ijmr.IJMR_2086_17.

[34] Vijayakumar S, Swetha RG, Bakthavatchalam YD, Vasudevan K, Abirami Shankar B, Kirubananthan A, et al. Genomic investigation unveils colistin resistance mechanism in carbapenem-resistant *Acinetobacter baumannii* clinical isolates. Microbiol Spectr 2024;12:e02511-23. https://doi.org/10.1128/spectrum.02511-23.

[35] Sharma S, Banerjee T, Yadav G, Kumar A. Susceptibility profile of blaOXA-23 and metallo-β-lactamases co-harbouring isolates of carbapenem resistant Acinetobacter baumannii (CRAB) against standard drugs and combinations. Front Cell Infect Microbiol 2023;12:1068840. https://doi.org/10.3389/fcimb.2022.1068840.

[36] Sharma S, Banerjee T, Yadav G, Palandurkar K. Mutations at Novel Sites in *pmrA/B* and *lpxA/D* Genes and Absence of Reduced Fitness in Colistin-Resistant *Acinetobacter baumannii* from a Tertiary Care Hospital, India. Microb Drug Resist 2021;27:628–36. https://doi.org/10.1089/mdr.2020.0023.
